# Critical interventions for demand generation in Zambia, Nepal, and Senegal with regards to the 5C psychological antecedents of vaccination

**DOI:** 10.1101/2022.04.25.22274035

**Authors:** Kyra A Hester, Zoe Sakas, Emily Awino Ogutu, Sameer Dixit, Anna S. Ellis, Chenmua Yang, Chama Chanda, Matthew C. Freeman, Walter A. Orenstein, Moussa Sarr, Robert A. Bednarczyk

## Abstract

**Introduction:** Childhood vaccination is an effective intervention for lowering the burden of infectious disease. Progress was made to increase coverage globally, but vaccine hesitancy and refusal has threatened to erode said increases. The 5C psychological antecedents of vaccination (“5C”) model provides a validated measure of “vaccine hesitancy or confidence” to assess individual thoughts and behaviors behind vaccination. Our purpose was to investigate population-level factors that contributed to high and sustained vaccination coverage via interventions in Zambia, Nepal, and Senegal, and alignment with the 5Cs.

**Methods:** FDGs and KIIs were collected at the national, regional, district, health facility, and community levels. We assessed the demand environment, as relayed by participants, and identified interventions that key informants reported as successful for demand generation, then retroactively aligned the interventions with the 5C constructs.

**Results:** Demand was positively correlated with high *confidence* and *collective responsibility*. Psychological *constraints* sometimes impacted demand. Physical *constraints* created barriers in some communities, particularly difficult to access (i.e., mountainous). Occasionally, physical *constraints* did not affect vaccination behavior - parents believed the benefits of vaccination worth pursuing. Factors negatively correlated with demand and intent, *complacency* and *calculation*, had limited impact. The most critical interventions were: targeted and tailored health education activities (i.e., media partnerships, school outreach); community engagement; community ownership; and involvement of community (i.e., community health workers, leaders, religious figures).

**Conclusion:** We found similar interventions were used to generate demand, and those strategies aligned with the 5C constructs. Categorizing interventions by drivers of demand may help strategic planning and the division of resources; decision makers may choose to implement our suggested interventions. Assessing the 5Cs allows for decision-makers to operationalize demand generation into concrete interventions and policies, and determine the individual impact of these constructs on the population and focus efforts on interventions tailored to a specific need.

## 1. Introduction

Reaching – and maintaining-high childhood vaccination coverage is one of the most cost-effective interventions to lower the burden of infectious disease. Globally, we have made progress toward increasing vaccine coverage; the third dose of diphtheria, tetanus, and pertussis (DTP3) vaccine among WHO member states increased from 72% in 2000 to 83% in 2020, but gaps remain [1-3]. Structural changes have been implemented to increase vaccination rates (e.g., cold chain expansion), but to ensure effective vaccine delivery, there needs to be active intent to vaccinate in addition to demand generation efforts by immunization programs.

Vaccine hesitancy and refusal to vaccinate has threatened to erode the gains in coverage [4, 5]. A 2013 survey of 13 immunization program managers from the 6 WHO Regions found that hesitancy was present across all countries; however, the impact of hesitancy on uptake varied by country-specific context [6]. The Strategic Advisory Group of Experts (SAGE) reviewed these findings, and determined that of countries implementing interventions to address hesitancy, “dialogue-based, directly targeted approaches can improve vaccine uptake, including engaging leaders, social mobilization, mass media, improving convenience, reminders, training for health-care workers, and increasing awareness”. Vaccine education for children was considered an area for further exploration [6].

The 5C psychological antecedents of vaccination (“5C”) model provides a validated measure of “vaccine hesitancy or confidence” that is used to assess individual thoughts and behaviors behind vaccination [7]. The five components of the 5C model include *confidence, complacency, constraints, calculation*, and *collective responsibility*, and is adapted from the SAGE Working Group on Vaccine Hesitancy’s original 3C model [5].The scale was validated through two studies; one to correlate psychological constructs with the scale, the other to correlate the scale with vaccination behavior. The resulting scale statistically correlates to vaccination behavior, and was predictive of future intent to vaccinate [5]. This model is considered ideal for assessing vaccination behavior [7-10].

The 5C model was developed with the intent to measure individual level behavior; it is currently unknown how this scale translates to community-level perceptions and activities, including how the government, programmatic, and community behaviors affect vaccination decision-making. Understanding the role of vaccine hesitancy at large may provide insight on the performance of demand generation activities. The purpose of this study was to review interventions that key informant interviewers (KIIs) attributed to increased vaccine uptake in Zambia, Nepal, and Senegal, and assess how these interventions affected the community vaccination behaviors as seen on the 5C model. We observed innovative approaches to generating demand as part of our research in Zambia, Nepal, and Senegal [11-13]. Interventions were often implemented from existing global guidelines and further refined to country context [14]. These interventions ultimately utilized constructs seen in the 5C model, and may be useful to other low- and middle-income countries (LICs and LMICs, respectively) within similar contexts to address challenges in vaccine uptake.

## 2. Methods

This multiple case study analysis was conducted using data from the Exemplars in Vaccine Delivery project within the Exemplars in Global Health program [15, 16]. We employed a qualitative analysis to investigate factors that contributed to high and sustained vaccination coverage through KIIs and focus group discussions (FGDs) at the national, regional, district, health facility, and community levels. We triangulated these findings with quantitative analyses using publicly available data.

Prior to data collection, we developed a conceptual model (Figure 1) to organize factors that impact childhood vaccine coverage globally. This model was based on the work of Phillips et al. and LaFond et al. alongside a broader review of the vaccine literature [17, 18]. While components of vaccine demand and intent are present in this model, we sought the use of the 5C model to further assess demand in our selected countries. The protocol of the primary analysis, along with individual case studies of exemplar countries and a synthesis case study are published separately [11-13, 15].

**Figure 1.**
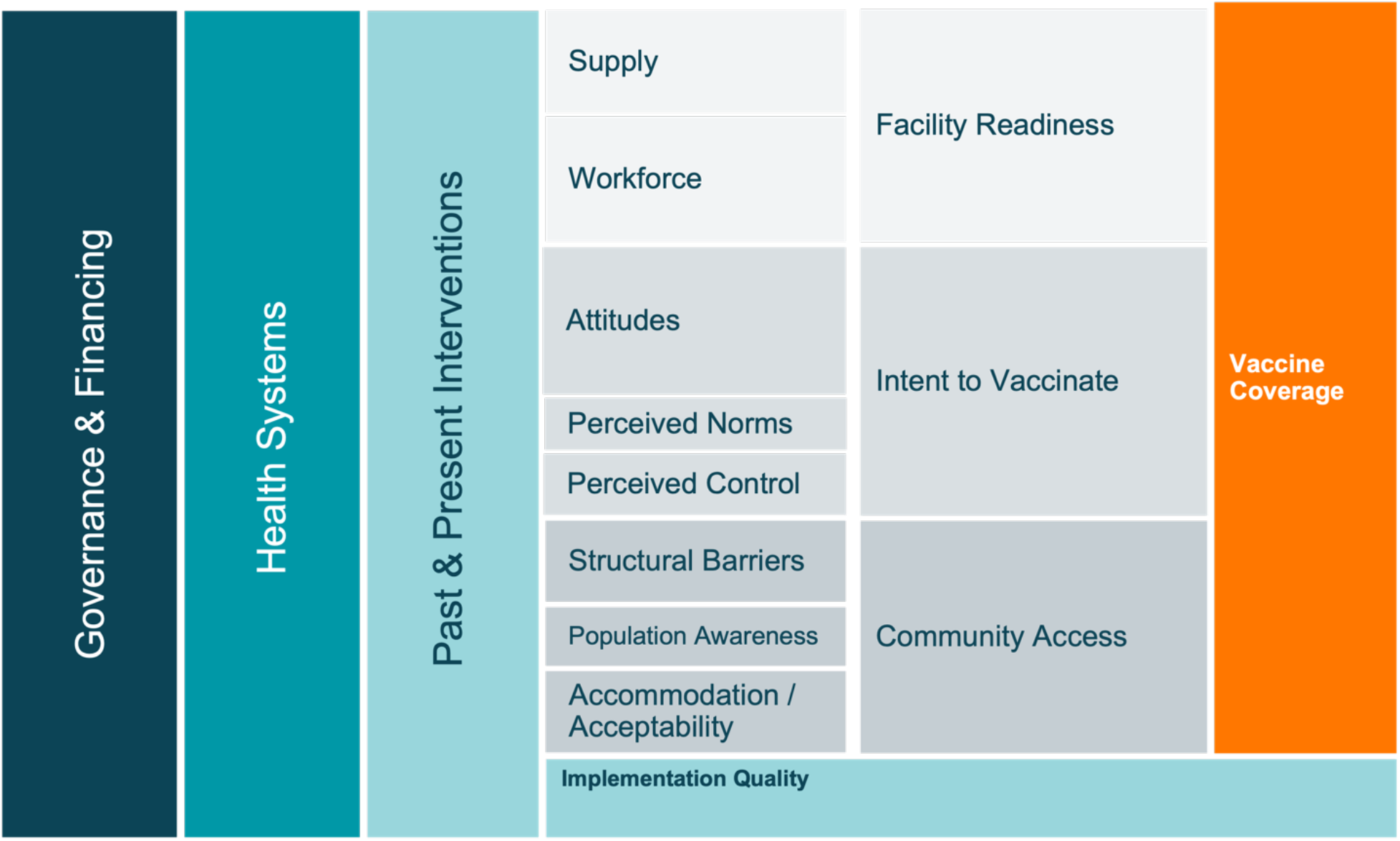
Conceptual model of the drivers of vaccine coverage.

### 2.1 Study Sites

Methods for country, region, and district selection are detailed elsewhere [11-13, 15]. Countries were selected based on DTP1 and DTP3 coverage estimates, which served as proxies for the evaluation of the vaccine delivery system in these countries, with DTP1 as a proxy of access and DTP3 as a proxy of continued utilization of immunization services (Figure 2) [15, 19]. Three regions within each country were identified in consultation with national stakeholders and available data (Table 2). Ministry of Health* officials facilitated site selection and data collection activities

**Table 1:**
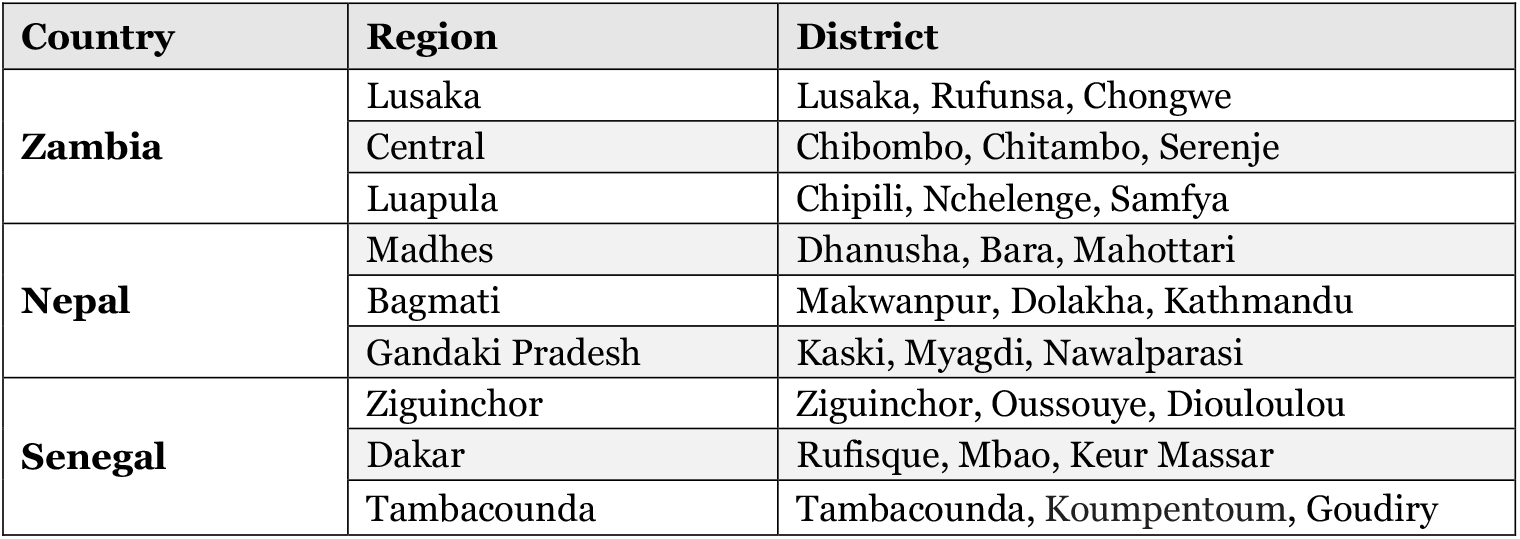
Summary of countries, regions, and districts selected for research.

**Table 2:**
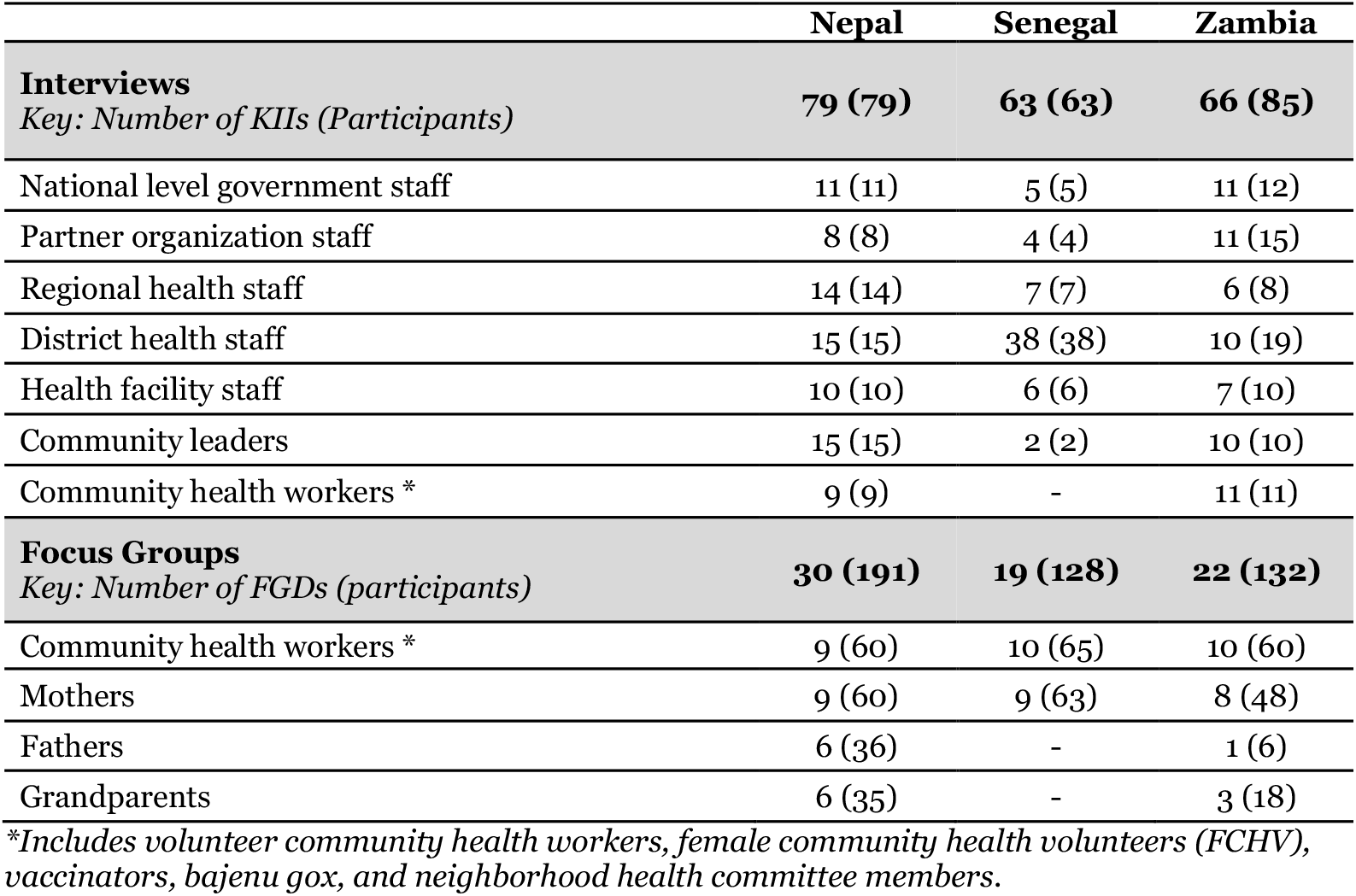
Summary of data collection activities.

|  | Nepal | Senegal | Zambia |
| --- | --- | --- | --- |
| **Interviews** |  |  |  |
| *Key: Number of KIIs (Participants)* | **79 (79)** | **63 (63)** | **66 (85)** |
| National level government staff | 11 (11) | 5 (5) | 11 (12) |
| Partner organization staff | 8 (8) | 4 (4) | 11 (15) |
| Regional health staff | 14 (14) | 7 (7) | 6 (8) |
| District health staff | 15 (15) | 38 (38) | 10 (19) |
| Health facility staff | 10 (10) | 6 (6) | 7 (10) |
| Community leaders | 15 (15) | 2 (2) | 10 (10) |
| Community health workers * | 9 (9) | - | 11 (11) |
| **Focus Groups** |  |  |  |
| *Key: Number of FGDs (participants)* | **30 (191)** | **19 (128)** | **22 (132)** |
| Community health workers * | 9 (60) | 10 (65) | 10 (60) |
| Mothers | 9 (60) | 9 (63) | 8 (48) |
| Fathers | 6 (36) | - | 1 (6) |
| Grandparents | 6 (35) | - | 3 (18) |

*\*Includes volunteer community health workers, female community health volunteers (FCHV), vaccinators, bajenu gox, and neighborhood health committee members.*

**Figure 2.**
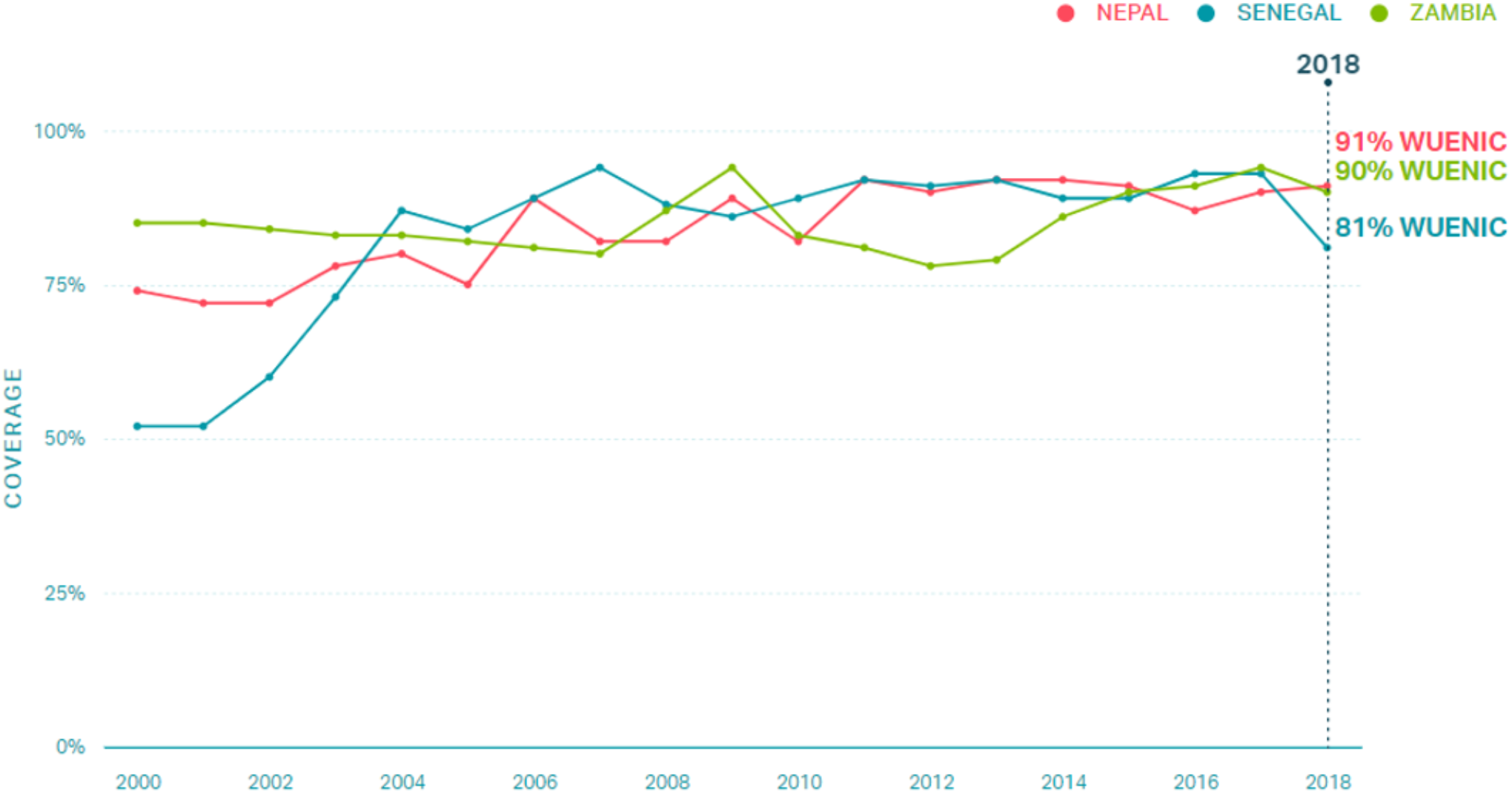
DTP3 coverage of Zambia, Nepal, and Senegal from 2000 to 2018; WUENIC data.

### 2.2 Qualitative Data collection and analysis

Qualitative data were collected between October 2019 and April 2021 at the national, regional, district, health facility, and community levels in Nepal, Senegal, and Zambia (Table 3). Interview guides were informed by the Consolidated Framework for Implementation Research (CFIR) and the Context and Implementation of Complex Interventions (CICI) frameworks [20]. KII and FGD guides were translated into local languages by research assistants. All interview guides were piloted before use and adjusted iteratively throughout data collection. An initial list of KIIs was developed with local research partners and MoH officials; snowball sampling was used to identify additional key informants. Our sampling approach included a diverse sample of participants in regard to geographic location and demographic qualities. Caregivers and volunteer community health workers (CHWs) were recruited for FGDs from health facility catchment areas with the assistance of local health staff. The duration of KIIs and FGDs averaged one and a half hours. KIIs and FGDs were audio-recorded with the permission of participants. Research files, recordings, and transcriptions were de-identified and password protected. We applied a theory-informed thematic analysis of the transcripts to identify critical success factors. Transcripts were coded and analyzed using MaxQDA2020 software (Berlin, Germany). Qualitative data collection and analysis tools, including topic guides and codebooks, can be found on our Open Sciences Framework (OSF) page [21].

**Table 3.**
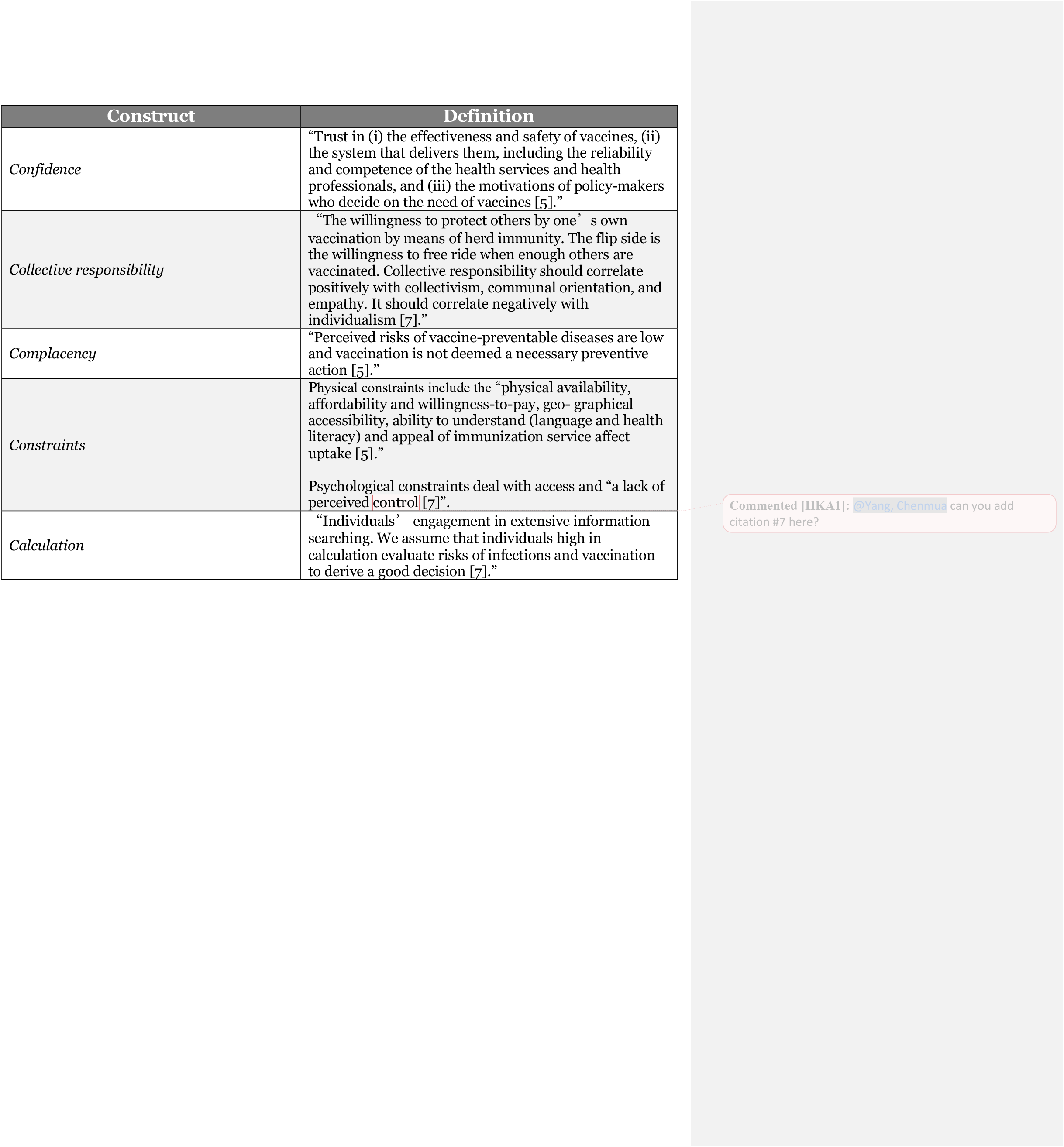
5C psychological antecedents of vaccination scale constructs and definitions.

### 2.3. Application of the 5C model

We analyzed qualitative data from Nepal, Senegal, and Zambia and pulled data coded with *public awareness, community engagement, media*, or *vaccine hesitancy*. We first assessed the current demand environment as relayed by KIIs and FGDs. Next, we identified interventions reported as most successful for demand generation and retroactively aligned the interventions with the constructs on the 5C model.

### 2.4. Ethical approvals

This study was considered exempt by the Institutional Review Board committee of Emory University, Atlanta, Georgia, USA (IRB00111474), and was approved by the Nepal Health Research Council (NHRC; Reg. no. 347/2019) in Kathmandu, Nepal; the National Ethical Committee for Health Research (CERNS; Comité National d’Ethique pour la Recherche en Santé) in Dakar, Senegal (00000174); the University of Zambia Biomedical Research Ethics Committee (Federal Assurance No. FWA00000338, REF. No. 166-2019); and the National Health Research Authority in Zambia. All participants provided written informed consent.

## 3. Results

We found that the 5C’s were an effective way to analyze intent and demand generation within our studied populations in Nepal, Senegal, and Zambia. Similar interventions across countries were used to generate demand, and these interventions ultimately aligned with all constructs of the 5c model (Table 4). Key informants from all three countries reported high levels of intent to vaccinate and demand for vaccines among their communities. Demand was positively correlated with high *confidence* and *collective responsibility*. Psychological *constraints* sometimes impacted demand for vaccine, hindering uptake. Physical *constraints* were barriers in some communities, particularly in those that were difficult to access (i.e., rural or mountainous communities). Occasionally, physical *constraints* did not affect vaccination behavior - parents believed the benefits of vaccination to be worth pursuing despite difficulties. Factors often negatively correlated with vaccine intent, such as *complacency* and *calculation*, were limited in impact in these three countries. Through interventions targeting health education and community ownership, the exemplar countries addressed community vaccination behavior within all constructs.

**Table 4.**
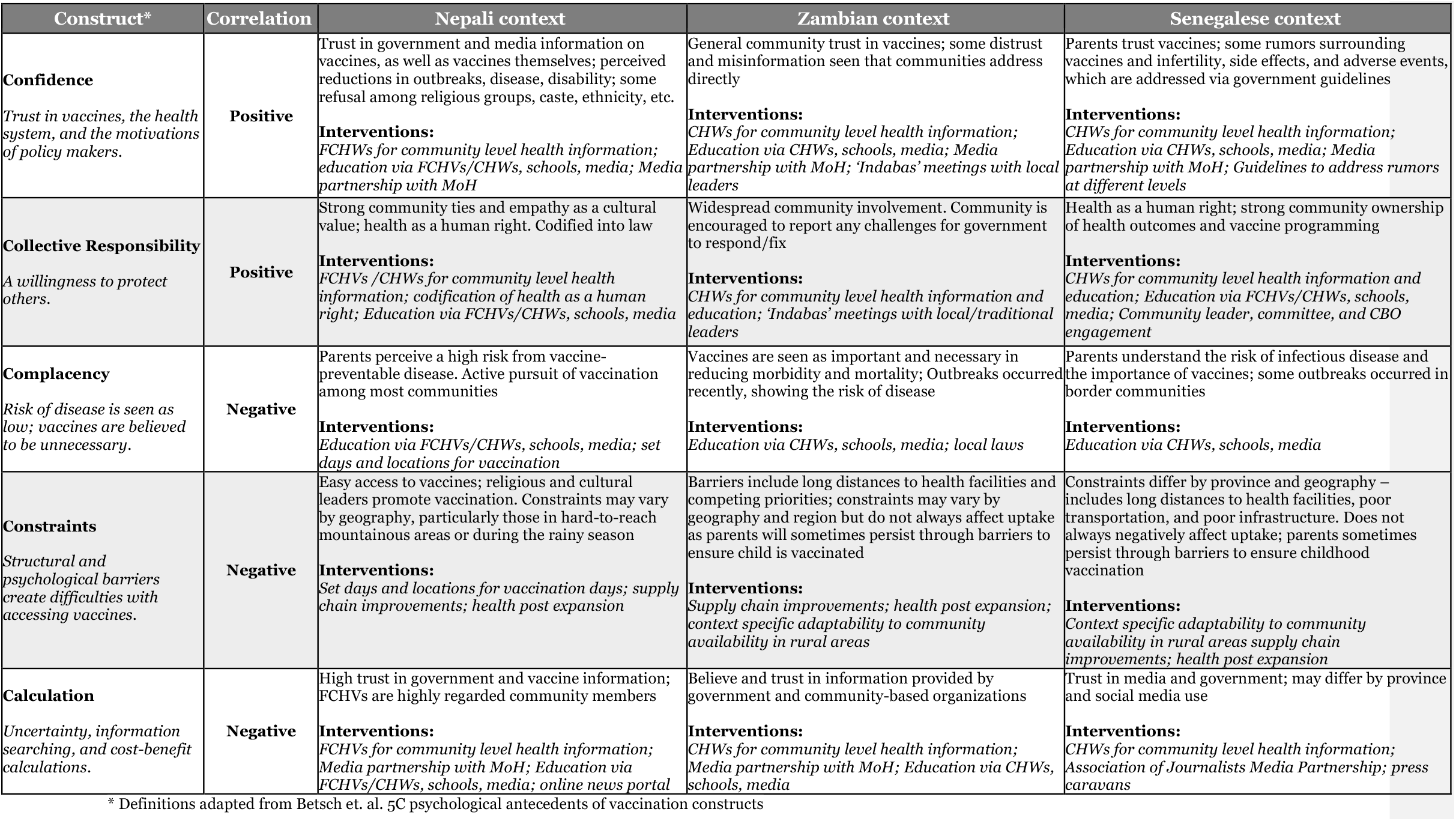
Application of the 5C model to Nepali, Zambian, and Senegalese context with the associated interventions utilized to raise demand.

### 3.1. Targeted and tailored health education activities

In all three countries, the MoH utilized existing channels – including media and schools – to disseminate factual information regarding vaccines. These targeted health education interventions aligned with each construct in the 5C model. *Confidence* in vaccines was fostered through dissemination of accurate and timely information from trusted sources, and raised the awareness of vaccine benefits. *Collective responsibility* was maintained by filtering vaccination messaging through societal norms and cultural ties. *Complacency* was addressed by education of parents and children on immunization and danger of infectious disease. The MoH addressed psychological *constraints* by providing media with tools to address misinformation and hesitancy along with information about upcoming vaccination events. *Calculation* was dissuaded by media and government transparency along with easy access to accurate information (Table 4).

#### Media partnerships

The MoH in Zambia, Nepal, and Senegal played an active role in engaging with the media for dissemination of vaccination information via strategic partnerships with media organizations and outlets. Within the MoH in all three countries, there were designated governmental agencies that handled media engagement. In Nepal, the National Health Education Information and Communication Center was responsible for providing media agencies with information for message development and verified or corrected information as needed. In Zambia, the Department of Health Promotions, Environmental Health, and Social Determinants provided the media with discussion points and scripts on new vaccine introductions, immunization campaigns, and general vaccinations. All countries had subnational offices for more tailored media dissemination; health workers at municipality levels referenced agreements between the community and media outlets (e.g., radio stations, TV, newspapers) to promote childhood vaccines and regularly announce routine vaccine days. National and local radio stations in Zambia provided free time slots for health messaging, and district and provincial health officers will go on air to discuss immunization activities.

In Senegal, the National Health Education and Information Service sits within the MoH and develops materials for health education nationally. The Association of Journalists for Health (L’Association des journalistes en santé, population et développement; AJSP) is an independent group of media experts that focuses on topics within the health and development sectors. According to key informants, the AJSP developed a “strategic partnership” with the MoH at the national level, and was composed of members from radio, TV, written press, and online media. The primary role of the AJSP was to ensure that accurate information about vaccines was disseminated to the public through multiple channels.

Countries utilized these partnerships to disseminate accurate information to communities, ultimately fostering trust in both the government and vaccines. At the local level, information was shared a variety of ways - radio being the most prevalent. Each country had unique modalities for media dissemination, such as an online news portal (Nepal), peer-to-peer learning from recorded visits (Zambia), and press caravans - defined as small scale campaigns (Senegal).

> *“They … deliver good messages to us. The information on the immunization month is discussed in every media like radio, local and national newspaper, online news portal, print, and by every level of the government. In case the regular program is delayed or not running, then they used to bring out these things in the media. So, the media has a positive role*.*” (District health official, Nepal)*

#### School outreach

The education systems in these countries were used to promote vaccination among children – either directly or indirectly. In Zambia, completed under-five cards - vaccination cards given at birth for recording vaccination status - were necessary in many communities for enrolling children in schools or registering for final exams. Although these requirements were not consistent, informants did report that under five cards fostered intent to vaccinate among parents. Schools hosted immunization weeks for children to catch-up on vaccinations, and teachers would provide immunization information to both parents and children.

Senegal and Nepal directly educated children on disease and vaccines. In both countries, the Ministry of Education (MoE) and MoH worked together to develop health education curriculums. The MoH in Senegal has collaborated with the MoE since the 1980s, and they produced courses on health and immunization for teachers, who share this information with school children as part of the National Education Program.

> *“In the initial teachers’ training, there is a module on immunization in the curriculum. There are courses that are given to explain [immunization]. That is why we ask the Ministry of Education to include their concerns about any health program in the curriculum in order to get the teachers to consider them as part of the teaching program. Malaria, for example, is part of the teaching program - as is vaccination. It is an aspect taken into account during the teacher’s evaluation*.*” (National health official, Senegal)*

In Nepal, the MoE and MoH collaborated through a formal partnership since 1971, when health was made a compulsory subject for all grade levels. The MoE developed a standard curriculum to influence the health knowledge, attitudes, and practices of students grades 1-10, taught to teachers during their training; only health workers approved by the MoHP could teach the topic [22, 23]. Teachers would educate their students about the date and place of immunization sessions in their communities, and would check the immunization status of at-risk populations.

> *“Yes, we have been doing school health programs. We have organized health-related topics in the curriculum of the education system while organizing health related programs and promoting good health behavior. We have organized training for students, teachers, the community and are reaching the community through school children. We organize these programs not only in terms of immunization, but all health-related programs and outbreak management. We made the format. This format has helped other health-related programs as well. We provide regular trainings and education every year*.*” (Regional health official, Nepal)*

### 3.2. Community engagement and ownership of health interventions and outcomes

Across all sites, respondents spoke to the importance of community actors for the implementation of demand generation activities – including their input on messaging and support for outreach. The MoH leveraged partnerships with community leaders, religious leaders, traditional leaders, and CHWs to increase vaccine demand and uptake. Interventions related to community ownership also aligned with the other antecedents for demand, by creating *confidence* in vaccines and service providers through the existing respect and trust of volunteer CHWs; addressing psychological *constraints* through tailored activities to local areas and individualized outreach to parents; addressing *complacency* by increasing public awareness through community channels; and limiting the impact of *calculation* through proactive engagement with community members and dissemination of accurate information (Table 4).

#### Community health workers

CHWs were influential players within the healthcare systems of Zambia, Nepal, and Senegal. In Nepal, Female Community Health Volunteers (FCHVs) were the main sources of vaccination information in communities. FCHVs are recruited to work in their own communities and utilize knowledge of community values, norms, and priorities to adjust messaging and personal communication to address the needs of specific groups. FCHVs ensure community members are knowledgeable of their right to receive vaccines and engage directly through door-to-door education. FCHV vaccine messaging has changed over time along with the needs of the community; initially, FCHVs informed parents to bring their children to vaccination days, without explaining why vaccinations are important. In recent years, FCHVs have focused more on thorough health education, including other long-term benefits of vaccines that impact quality of life.

> *“Previously, the people were only informed to bring their children for vaccination, but now we say why it is important to vaccinate. When they ask we link it with financial things and also say the investment must be done in health*.*” (District health official, Nepal)*

In Zambia and Senegal, CHWs provided tailored messaging to communities to encourage and reinforce vaccination. In Senegal, CHW cadres called *relais* and *bajenu gox* (“godmothers”) were critical to the success of the vaccination program. They sensitized communities several days before vaccine days and checked vaccination cards to assess whether children needed to attend upcoming events. In some cases, they called or texted parents to remind them of vaccination days. These CHWs provided extensive follow-up on dropouts, including door-to-door activities, and encouraged reluctant parents to vaccinate their children.

> *“The bajenu gox come to check if we are respecting the vaccination calendar, and if they notice any irregularities when they consult our booklet they ask us to go and catch up*.*” (Mother, Senegal)*

Zambia utilizes Neighborhood Health Committees (NHCs) to dispel myths and misconceptions about vaccinations. NHCs are community-appointed volunteers whose primary role is to coordinate and supervise other Community Based Volunteers on behalf of the health facility staff; NHCs serve as the link between the health system and community [24]. Since NHCs are recruited by community leaders and work closely with leaders and MoH staff, they are a trusted source of information. NHCs also help coordinate general health service delivery, including community-level immunization services.

> *I think the myths and misconceptions was tackled through the interaction we were having with the NHCs. We came up with a program of meeting with the NHCs every quarter, and meeting - we call them the “chilolo’s”, the headmen - twice per year to explain the importance of vaccines*.*” (District health official, Zambia)*

#### Community leaders, organizations, and religious figures

Dissemination through community leaders proved impactful in all three countries as communities trusted the information source. Involvement from community leaders and community-based organizations were seen as critical to connect communities to the healthcare system, and community actors acted as gatekeepers to their communities. In Senegal, supportive community actors, including chiefs, village opinion leaders, neighborhood delegates, traditional birth attendants, marabouts (traditional doctors), and religious leaders (e.g., imams) conducted vaccination information sessions and followed-up with reluctant parents. Nepal has a similar type of community engagement, with traditional leaders, local leaders, FCHVs, religious figures, and teachers providing counsel and encouragement to parents. Community-led groups are present in both Senegal and Nepal, and are involved with immunization programming. This included promotion, service delivery, and targeted outreach, in addition to supporting equity and inclusivity by involving members from a variety of religious, ethnic, and socioeconomic groups.

> *We also involve religious leaders, as you say. The neighborhood delegates get involved in any case. It is all the leaders who can really help us to strengthen this communication and to raise awareness in relation to vaccination. So, it is according to the context of each area of responsibility, but everyone is involved - from the delegates to the mayors*… *There are a lot of strategies that are implemented in the districts, to reach the maximum of the community. (District health official, Senegal)*

In Nepal, mothers are also connected to the health system through Mothers’ Health Groups (MHGs). MHGs are formed with the involvement of the community, local health institutions, and local governments, and consist of “all interested mothers of reproductive age” [25]. A discussion topic is chosen as a group, with one FCHV facilitating. MHGs meet monthly and discuss various health topics, including immunizations. FCHVs conducted these meetings once a month; often on the same day as the immunization or outreach clinic [26]. Women in MHGs are tasked with sharing information from the meetings with other mothers in the community.

> *Basically, the Mothers’ Group is responsible for giving education and information to the parents. We give and focus education and information on the mother rather than the father. If the mother knows about immunization camp, then the mother takes their baby in any condition or situation. FCHVs and Mothers’ Group provide information regarding immunization activities or regular immunization programming. For new vaccines, it is discussed with FCHVs and mothers’ group because, unless they support and are involved in the program, it won’t be successful because they are the ones who are aware, give knowledge and information to the people at the grassroot level. (Former national level official, Nepal)*

MoH officials in Zambia hold *indabas*, or formal community meetings, at least once per year at the provincial level. These meetings include all traditional and religious leaders in the area. The *indabas* were created in 2010 to educate community leaders on community health – including vaccinations – and to distribute relevant information to the community. Health facility staff meet with community leaders on an ad hoc basis; usually every 1-3 months. Leaders participate and have the opportunity to share challenges faced in their community, and suggest solutions. Meetings may also include planning and communication for upcoming activities (including Child Health Week), campaigns for the introduction of new vaccines, and notification of disease outbreaks. Upon completion of the *indabas*, the ministry and community leaders develop a plan to address health issues in their communities. Zambian community leaders then share information learned from *indabas* with their constituents. In addition, Zambia has been particularly successful at leveraging social civil society organizations like the Churches Health Association of Zambia to help increase trust and participation in the immunization program [27]. These organizations assist by providing information about vaccination programs and dispelling myths.

> *“At some point, we’ve even had some provincial Indabas where all the traditional leaders from almost all the districts are called to attend these meetings. So, it’s not something that is starting this year or started last year because I remember even as far back as 2018 we had a meeting…. For key traditional leaders from across the province we meet with them …. in Mansa to discuss some of these issues. I know there were follow up meetings in the districts so it’s not starting now, it has always been a priority*.*” (District health official, Zambia)*

## 4. Discussion

We conducted a qualitative study to assess interventions attributed to increased vaccine coverage by KIIs in Zambia, Nepal, and Senegal; the 5C model was then utilized to analyze the impact of the interventions on community vaccination behaviors. Our novel approach applied the 5C model by assessing the constructs at a population, rather than individual, level. This population level assessment of intent and demand for vaccine ultimately correlated to the model, illustrating that further use of the 5C’s may be useful for country-level stakeholders to determine wide-scale facilitators and barriers to vaccination.

We found *collective responsibility* to be the construct most correlated to demand generation in these countries. All studied countries exhibited cultural norms that related to *collective responsibility*, with participants from the community and health sector alike stressing the importance of the collective health of their communities. Vaccinations were considered critical to children’s health by these populations, and in Nepal and Senegal, laws were codified to ensure all have access to vaccines; through codification, the governments legally confirmed the importance of community health and their responsibility in ensuring access [12, 13]. Although *collective responsibility* was the most prominent construct seen across the countries, interventions that utilized this construct may be the most difficult to replicate due to differences in culture and values. While ultimately similar, these community values arose through different means in Nepal and Senegal – but both countries are known to have strong social cohesion [28-31]. Countries with ‘tighter’ social cohesion were found to experience a ‘tipping point’ for norm change, compared to countries with ‘looser’ social cohesion that shows gradual change. This was seen during the COVID-19 pandemic, and Gelfand et al determined that countries with cultural ‘looseness’ had about 5 times the cases and 8.75 times the deaths than countries who had cultural ‘tightness’ and “abide by strict norms” [32]. Government initiatives from Zambia, Nepal, and Senegal focused on community engagement, involvement of local leaders, and targeted community messaging to operationalize existing social values and cohesion and to further increase intent and demand for vaccines.

*Confidence* in the safety and efficacy of vaccines, providers, and the government at large fostered demand for vaccines in all three countries. Trust in vaccine efficacy was achieved through tangible decreases in morbidity and mortality witnessed by older generations who remembered the amount of disease present before vaccines were widely available. In Senegal, elderly women were specifically recruited for public awareness activities due to their unique and valuable perspectives. All three countries relied on trusted, volunteer, CHWs to improve *confidence* in service delivery by acting as a bridge to connect health staff to the people living in their catchment areas. CHWs are recognized as critical connections to the community, and provide culturally sensitive care and communication [33, 34]. When appropriately trained and supervised, CHWs in both LMIC and HIC contexts have been associated with improved health outcomes and accessibility to services [35]. In addition to primary health education activities, they are mobilized to generate health service demand within underserved, low-income, indigenous, and marginalized populations [34-37]. Japan’s government plays a leading role to recruit, train and recognize CHWs in the healthcare system [38]. However, due to a lack of formal training, CHWs in other HICs are often recruited on an ad-hoc basis by community-based organizations and trained on-the-job as temporary solutions to address health disparities [35, 36]. Similar to Senegal, elderly CHWs in Japan are specifically recruited for their homogeneity, trustworthiness, and years of experience in a community to increase overall involvement and effect of health promotion activities [39, 40]. CHWs can be utilized to reach populations facing vaccination barriers and increase rates [41, 42]. One recent study identified potential benefits of CHW-led interventions to increase influenza vaccination awareness among migrants and refugees in the United States 43]. The authors found multicomponent CHW-led interventions can increase vaccination knowledge and understanding, but further supports that health education must be provided by trusted community health members/CHWs and packaged with improved access to improve uptake [43]. Ultimately, these studies of CHWs in HICs show the utility in application of our findings throughout a multitude of settings.

Not all constructs were directly transferrable at a population level as currently described in the 5C model. For example, population level demand had improved alignment with *constraints* once we separated it into two subcategories – physical *constraints* and psychological *constraints*. When looking at physical *constraints* separately, we determined that for some communities, access did not affect demand; in fact, parents’ demand for vaccines was high enough to make physical barriers, such as traveling long distances or waiting long periods, worth pressing through to protect their children from diseases. We theorize this difference may be explained by the LMIC status of Nepal, Senegal, and Zambia, and recommend that further use of the 5C model in LIC/LMIC settings have the constraint construct separated into these distinct sub-categories. Additionally, we hypothesize that communities with recent outbreaks or generational memory of vaccine preventable disease may make the choice to vaccinate regardless of physical *constraints*. Despite these differences, interventions targeted to 5C model constructs may provide ministries, program managers, and/or donors with evidence-based ways to increase demand - particularly in LICs/LMICs.

### 4.1 Application of our findings

Our findings highlight that a population level assessment of the constructs in the 5C model can guide decision-makers in the creation - or adaptation - of interventions to increase intent and demand for childhood vaccines. Categorizing interventions by what drives demand may support strategic planning and influence the division of resources; policy makers may opt to ensure that demand generation activities cover all five constructs - increasing *confidence* and *collective responsibility* and addressing *complacency, constraints*, and *calculation*. Decision-makers may also use this assessment strategy to determine the individual impact of these constructs on the population, and focus their efforts on interventions that are tailored to a specific need. In Nepal, Senegal, and Zambia, we found that all five constructs were addressed through targeted education, media partnerships, CHW engagement, and buy-in from community and religious leaders - leading to generally high levels of demand, and ultimately vaccination coverage. From our work in Zambia, Nepal, and Senegal, we recommend the following strategies to target all 5C constructs and improve population-level demand:

1. Utilize existing community structures and consider cultural and social norms when designing interventions.
2. Disseminate vaccine information through multiple trusted sources, such as CHWs, newspapers, radios, and religious leaders.
3. Conduct school outreach by providing schoolteachers with lesson plans on health, including the importance of vaccination.
4. Assess physical and psychological constraints separately in LIC/LMIC settings and respond to each via tailored programming.

## 5. Limitations

There are several limitations to this study. First, the 5C model was created, validated, and mostly utilized, within HICs. Our research in LMICs mapped clearly to the model, which may make this applicable to population and systems levels; this is something that should be explored. Third, all countries had a cultural norm surrounding *collective responsibility*, therefore, these identified interventions may work best in similar cultures.

## 6. Conclusion

Demand for childhood vaccines was high among community members in countries with high and sustained coverage. Interventions used to generate demand –via raising public awareness and fostering community ownership of health– mapped onto the existing 5C psychological antecedents of vaccination framework. Utilizing theory-driven strategies like the 5C model may provide structure to community demand generation via identification of areas where additional interventions are needed to improve intent and demand. Interventions within Nepal, Senegal, and Zambia demonstrated fostering *confidence* and *collective responsibility* through a culture of community, while addressing barriers like physical and psychological *constraints, complacency*, and *calculation* with targeted education and messaging, alongside active community engagement.

## Data Availability

De-identified data are available upon request.

https://osf.io/7ys4a/?view_only=739ca7a72f9749118b4aa3d2f7b655d9

## Acknowledgements

We thank the Center for Family Health Research in Zambia, the Center for Molecular Dynamics Nepal, along with Institut de Recherche en Santé de Surveillance Epidemiologique et de Formation in Dakar, Senegal. We gratefully acknowledge the participants who gave their time and insights to help us better understand the vaccine delivery systems of Zambia, Nepal, and Senegal, along with facilitators from their respective Ministries of Health.

## Data statement

De-identified data are available upon request.

## Footnotes

* *Note that each country has a slightly different name for their Ministry of Health (e*.*g*., *Ministry of Health and Population in Nepal; Ministry of Health and Social Action in Senegal), but all will be referred to as “MoH” throughout for simplicity*.

## References

1. Hwang, A., et al., Global Vaccine Action Plan Lessons Learned II: Stakeholder Perspectives. Vaccine, 2020. 38(33): p. 5372–5378.

2. Cherian, T., et al., Global Vaccine Action Plan lessons learned III: Monitoring and evaluation/accountability framework. Vaccine, 2020. 38(33): p. 5379–5383.

3. World Health Organization, Diptheria tetanus toxoid and pertussis (DTP3) immunization coverage among 1-year-olds (%). 2021, World Health Organization: Geneva.

4. Cooper, S., et al., Vaccine hesitancy – a potential threat to the achievements of vaccination programmes in Africa. Human Vaccines & Immunotherapeutics, 2018. 14(10): p. 2355–2357.

5. MacDonald, N. and the SAGE Working Group on Vaccine Hesistancy, Vaccine hesitancy: Definition, scope and determinants. 2015.

6. World Health Organization, Meeting of the Strategic Advisory Group of Experts on immunization, October 2014 — conclusions and recommendations. 2014. p. 561–576.

7. Betsch, C., et al., Beyond confidence: Development of a measure assessing the 5C psychological antecedents of vaccination. PLOS ONE, 2018. 13(12): p. e0208601.

8. Machida, M., et al., Trends in COVID-19 vaccination intent from pre-to post-COVID-19 vaccine distribution and their associations with the 5C psychological antecedents of vaccination by sex and age in Japan. Human Vaccines & Immunotherapeutics, 2021. 17(11): p. 3954–3962.

9. Hossain, M.B., et al., Health Belief Model, Theory of Planned Behavior, or Psychological Antecedents: What Predicts COVID-19 Vaccine Hesitancy Better Among the Bangladeshi Adults? Frontiers in Public Health, 2021. 9.

10. Nath, R., et al., Role of Vaccine Hesitancy, eHealth Literacy, and Vaccine Literacy in 4Young Adults&rsq o; COVID-19 Vaccine Uptake Intention in a Lower-Middle-Income 475 Country. Vaccines, 2021. 9(12): p. 1405.

11. Rodriguez, K., et al., Critical success factors for routine immunization performance: A case study of Zambia 2000 to 2018. medRxiv, 2021: p. 2021.11.30.21267060.

12. Sakas, Z., et al., Critical success factors for high routine immunization performance: A case study of Senegal. medRxiv, 2022: p. 2022.01.25.22269847.

13. Hester, K.A., et al., Critical success factors for high routine immunization performance: A case study of Nepal. medRxiv, 2022: p. 2022.01.28.22270023.

14. WHO, Global Vaccine Action Plan 2011-2020. 2013, World Health Organization: Geneva, Switzerland.

15. Bednarczyk, R.A., et al., Protocol: Identification and evaluation of critical factors in achieving high and sustained childhood immunization coverage in selected low-and lower-middle income countries. medRxiv, 2021: p. 2021.12.01.21267018.

16. Exemplars in Global Health. Making Better Decisions in Global Health: Understand Positive Outliers to Inform Policy and Practice. 2021; Available from: https://www.exemplars.health/.

17. Phillips, D.E., et al., Determinants of effective vaccine coverage in low and middle-income countries: a systematic review and interpretive synthesis. BMC health services research, 2017. 17(1): p. 681–681.

18. LaFond, A., et al., Drivers of routine immunization coverage improvement in Africa: findings from district-level case studies. Health policy and planning, 2015. 30(3): p. 298–308.

19. Nepal Demographic Health Survey 2001, 2006, 2011, 2016, 2017, DHS Program, Editor.

20. Pfadenhauer, L.M., et al., Making sense of complexity in context and implementation: the Context and Implementation of Complex Interventions (CICI) framework. Implementation science: IS, 2017. 12(1): p. 21–21.

21. Hester K, Open Sciences Framework (OSF) page: Exemplars in Vaccine Delivery. https://osf.io/7ys4a/?view_only=739ca7a72f9749118b4aa3d2f7b655d9

22. Sthapit, N.M., An inservice health education curriculum for primary teachers in Nepal.. 1979.

23. Nepal Ministry of Education and Sports, Primary Education Curriculum 2063: Grade 1-3. 2007.

24. Republic of Zambia Ministry of Health, Zambia National Health Strategic Plan 2017-2021.

25. Government of Nepal Ministry of Health and Population, National Female Community Health Volunteer Program Strategy, Family Health Division, Editor. 2007: Nepal.

26. Rajbhandari S H.S., Sanghvi H, McPherson R, Pradhan YV, Baqui AH, Expanding uterotonic protection following childbirth through community-based distribution of misoprostol: Operations research study in Nepal. Int J Gynaecol Obstet, 2009.

27. Churches Health Association of Zambia. Churches Health Association of Zambia: About CHAZ. Available from: https://www.chaz.org.zm/about-chaz/.

28. Gilligan, M.J., B.J. Pasquale, and C. Samii, Civil War and Social Cohesion: Lab-in-the-Field Evidence from Nepal. American Journal of Political Science, 2014. 58(3): p. 604–619.

29. Munroe, R.L., Altruism and Collectivism: An Exploratory Study in Four Cultures. Cross-Cultural Research, 2017. 52(3): p. 334–345.

30. Wilfahrt, M., The politics of local government performance: Elite cohesion and cross-village constraints in decentralized Senegal. World Development, 2018. 103: p. 149–161.

31. De, S., et al. Tipping Points for Norm Change in Human Cultures. in Social, Cultural, and Behavioral Modeling. 2018. Cham: Springer International Publishing.

32. Gelfand, M., et al., The relationship between cultural tightness-looseness and COVID-19 cases and deaths: a global analysis. The Lancet Planetary Health, 2021. 5.

33. Glenton, C., D. Javadi, and H.B. Perry, Community health workers at the dawn of a new era: 5. Roles and tasks. Health Research Policy and Systems, 2021. 19(3): p. 128.

34. Javanparast, S., et al., Community Health Worker Programs to Improve Healthcare Access and Equity: Are They Only Relevant to Low-and Middle-Income Countries? International journal of health policy and management, 2018. 7(10): p. 943–954.

35. Perry, H.B., R. Zulliger, and M.M. Rogers, Community Health Workers in Low-, Middle-, and High-Income Countries: An Overview of Their History, Recent Evolution, and Current Effectiveness. Annual Review of Public Health, 2014. 35(1): p. 399–421.

36. Najafizada, S.A.M.M.D., et al., Community health workers in Canada and other high-income countries: A scoping review and research gaps. Canadian Journal of Public Health, 2015. 106(3): p. E157–E164.

37. Ospina, J.E., et al., Community health workers improve contact tracing among immigrants with tuberculosis in Barcelona. BMC Public Health, 2012. 12(1): p. 158.

38. Taguchi, A., et al., Recruiting, trianing, and supporting community based health promotion volunteers in Japan: findings from a national survey, in 146th American Public Health Association Annual Meeting & Exposition 2018: San Diego. p. 10–14.

39. Murayama, H., A. Taguchi, and S. Murashima, Differences in Psychosocial Factors Among Novice, Experienced, and Veteran Health Promotion Volunteers in Japan. Public Health Nursing, 2008. 25(3): p. 253–260.

40. Murayama, H., A. Taguchi, and S. Murashima, Does Similarity in Educational Level Between Health Promotion Volunteers and Local Residents Affect Activity Involvement of the Volunteers? Public Health Nursing, 2012. 29(1): p. 36–43.

41. Malik, M.N., M.S. Awan, and T. Saleem, Social mobilization campaign to tackle immunization hesitancy in Sargodha and Khushab districts of Pakistan. Journal of global 550 health, 2020. 10(2): p. 021302–021302.

42. Rahman, R., A. Ross, and R. Pinto, The critical importance of community health workers as first responders to COVID-19 in USA. Health promotion international, 2021. 36(5): p. 1498–1507.

43. Ponce-Gonzalez, I.M., et al., A Multicomponent Health Education Campaign Led by Community Health Workers to Increase Influenza Vaccination among Migrants and Refugees. Journal of primary care & community health, 2021. 12: p. 21501327211055627–21501327211055627.

